# Comparison of renin–angiotensin–aldosterone system inhibitors with other antihypertensives in association with coronavirus disease-19 clinical outcomes: systematic review and meta-analysis

**DOI:** 10.1101/2020.05.21.20108993

**Authors:** Yihienew M. Bezabih, Alemayehu Bezabih, Endalkachew Aalamneh, Gregory M. Peterson, Woldesellassie M. Bezabhe

## Abstract

**Introduction:** The effects of renin–angiotensin–aldosterone system (RAAS) inhibitors on the clinical outcomes of coronavirus disease-19 (COVID-19) have been conflicting in different studies. This meta-analysis was undertaken to provide more conclusive evidence.

**Methods:** A systematic search for published articles was performed in PubMed and EMBASE from January 5 2020 till May 5 2020. Studies that reported the clinical outcomes of patients with COVID-19, stratified by the class of concomitant antihypertensive drug therapy, were included. The Mantel-Haenszel random effects model was used to estimate pooled odds ratio (OR).

**Results:** A total of 6,997 patients with COVID-19 were included, and all of them had hypertension. The overall risk of poor patient outcomes (severe COVID-19 or death) was lower in patients taking RAAS inhibitors (OR=0.84, 95% CI: [0.73, 0.96]; P=0.017) compared with those receiving non-RAAS inhibitor antihypertensives. Patients taking angiotensin-I-converting enzyme inhibitors (ACEIs) were less likely to experience poor clinical outcomes (OR=0.73, 95% CI: [0.58-0.92]; P=0.01) compared with those receiving angiotensin-II receptor blockers (ARBs). In addition, comparison of ACEIs to the rest of non-ACEI antihypertensives gave a consistently decreased risk of poor COVID-19 outcome (OR=0.77, 95% CI: [0.63-0.93]; P=0.002). However, ARBs did not decrease the risk of poor COVID-19 outcomes compared to all other non-ARB antihypertensives (OR=1.13, 95% CI: [0.95-1.35]).

**Conclusion:** The risk of developing severe illness or death from COVID-19 was lower in patients who received RAAS inhibitors compared with those who took non-RAAS inhibitors. ACEIs might be better in decreasing the severity and mortality of COVID-19 than ARBs.

## Introduction

The effect of renin–angiotensin–aldosterone system (RAAS) inhibitors on the clinical outcomes of coronavirus disease-19 (COVID-19) is of great interest ^1^. This is because RAAS blockers, one of the most commonly prescribed antihypertensive drug groups, were previously reported to have some interactions with the pathophysiology of severe acute respiratory syndrome coronavirus 2 (SARS-CoV-2) ^1,2^

Experimental studies have shown that, blockage of RAAS by either angiotensin-I-converting enzyme inhibitors (ACEIs) or angiotensin-II receptor blockers (ARBs) substantially upregulates the expression of host angiotensin-converting enzyme 2 (ACE_2_)^3^, a transmembrane enzyme used by SARS-CoV-2 as a receptor to enter and infect cells ^4^ On the other hand, ACE_2_ catalyzes the degradation of potentially harmful angiotensin-II to a vasodilator angiotensin (1-7), which has antiarrhythmic and cardioprotective effects^3,2^. In addition, RAAS inhibitors may also prevent some complications of COVID-19, such as hypokalaemia. Hence, despite concerns that overexpression of ACE_2_ with RAAS inhibitors could facilitate infection of tissues by SARS-CoV-2, these drugs could also have a therapeutic role.

Recent studies on the effects of RAAS inhibitors (ACEIs and ARBs) on the clinical outcomes of patients with COVID-19 have reported conflicting results, ranging from a decrease in mortality^5,6^, no effect^7–11^ or even an increase in mortality^12^. Therefore, our primary objective was to perform a systematic review and meta-analysis to estimate the overall risk of poor COVID-19 outcomes in patients receiving RAAS inhibitors compared to non-RAAS inhibitor antihypertensive agents. As secondary objective, we also compared the of risk developing poor clinical outcomes between specific classes of antihypertensives (ACEIs, ARBs, beta-blockers (BBs), calcium channel blockers (CCBs), and thiazides).

## Methods

This study was conducted following the Preferred Reporting Items for Systematic Reviews and Meta-Analyses (PRISMA) 2009 checklist^13^.

### Data sources and search terms

We searched PubMed and EMBASE to identify potentially relevant articles published between January 5 2020 to May 5 2020. A grey literature search was also performed to find additional articles that may have not been indexed. We used three main search keywords: (1) clinical outcome OR death OR mortality, (2) angiotensin and (3) COVID. These key words were combined with Boolean operators to make the following search term: (((((clinical outcome) OR death) OR mortality)) AND angiotensin) AND COVID. We found 53 and 64 articles indexed in PubMed and EMBASE, respectively. One additional article was found from a manual search. Two authors (Y. B., W. B.) selected studies by screening titles and abstracts. A third author (E. A) served as a mediator to reach a consensus for discrepancies.

### Study definitions

RAAS inhibitors in this study refer to only ACEIs and ARBs. Severe COVID-19 refers to the presence of any of the following: respiratory rate ≥30 breaths/minute, oxygen saturation at rest ≤93%, oxygenation index [PaO2/FiO2] ≤300 mm Hg, respiratory or other organ failure, mechanical ventilation, shock, or intensive care unit treatment^14^. We used the term ‘poor clinical outcome’ to indicate the presence of either severe COVID-19 or death.

### Outcome of Interest

The main outcome of interest was the risk of having poor clinical outcomes in patients infected with COVID-19 while receiving RAAS inhibitors, compared with those taking other antihypertensive agents. The secondary outcome was the risk of severe COVID-19 or death in patients receiving ACEIs inhibitors compared with those receiving ARBs or other classes of antihypertensives.

### Study selection: inclusion and exclusion criteria

Studies that reported the clinical outcomes of COVID-19 patients stratified by class of antihypertensive drug therapy for a similar comorbidity were included. Cohort (prospective or retrospective) studies, case series and editorials/letters that assessed COVID-19 clinical outcomes for patients taking RAAS inhibitors versus non-RAAS inhibitors were included. The included papers were either published or accepted original articles written in English. We excluded review papers and case reports. In addition, studies that examined COVID-19 clinical outcomes in heart failure patients on RAAS inhibitors were ineligible. This is because ACEs and ARBs are generally given to heart failure patients when the disease is worse (ejection fraction <40%) for the prevention of cardiac remodelling^15^.

### Data extraction and quality control

In each study, the total number of patients taking RAAS inhibitors or other class(es) of antihypertensives was recorded. Then, for each antihypertensive class exposure, the total number of patients with a poor clinical outcome (severe COVID-19 or death) versus those with a good outcome (non-severe COVID-19 and survival) were recorded. In addition, year, design of study and nature of comorbidities were also documented (Table 2).

The Newcastle-Ottawa quality assessment scale (NOS)^16^ was used for quality assessment of the included studies (Supplementary Table S1). Two reviewers (W.B. and E.A.) independently performed the quality assessment and another author (Y.B.) brought consensus during discrepancies. Articles which got a score of less than 7 stars in the NOS were considered poor quality and excluded (Supplementary Table 1).

### Data Analysis

The Mantel-Haenszel random effects model was used to estimate pooled odds ratio (OR), and a two-side alpha value less than 0.05 was considered significant. Publication bias was assessed using the funnel plot asymmetry. All the analysis were performed using the OpenMeta (Analyst) ^17^

## Results

### Study characteristics and quality assessment

A total of 117 potentially relevant articles through EMBASE (64), PubMed (53), and manual search (1) were found. Of these, 7 articles were included in our final analysis (Figure 1). All the included articles were of good quality (NOS score ≥7), and study characteristics and quality assessment are shown in Table 1 and Supplementary table S1, respectively.

**Figure 1:**
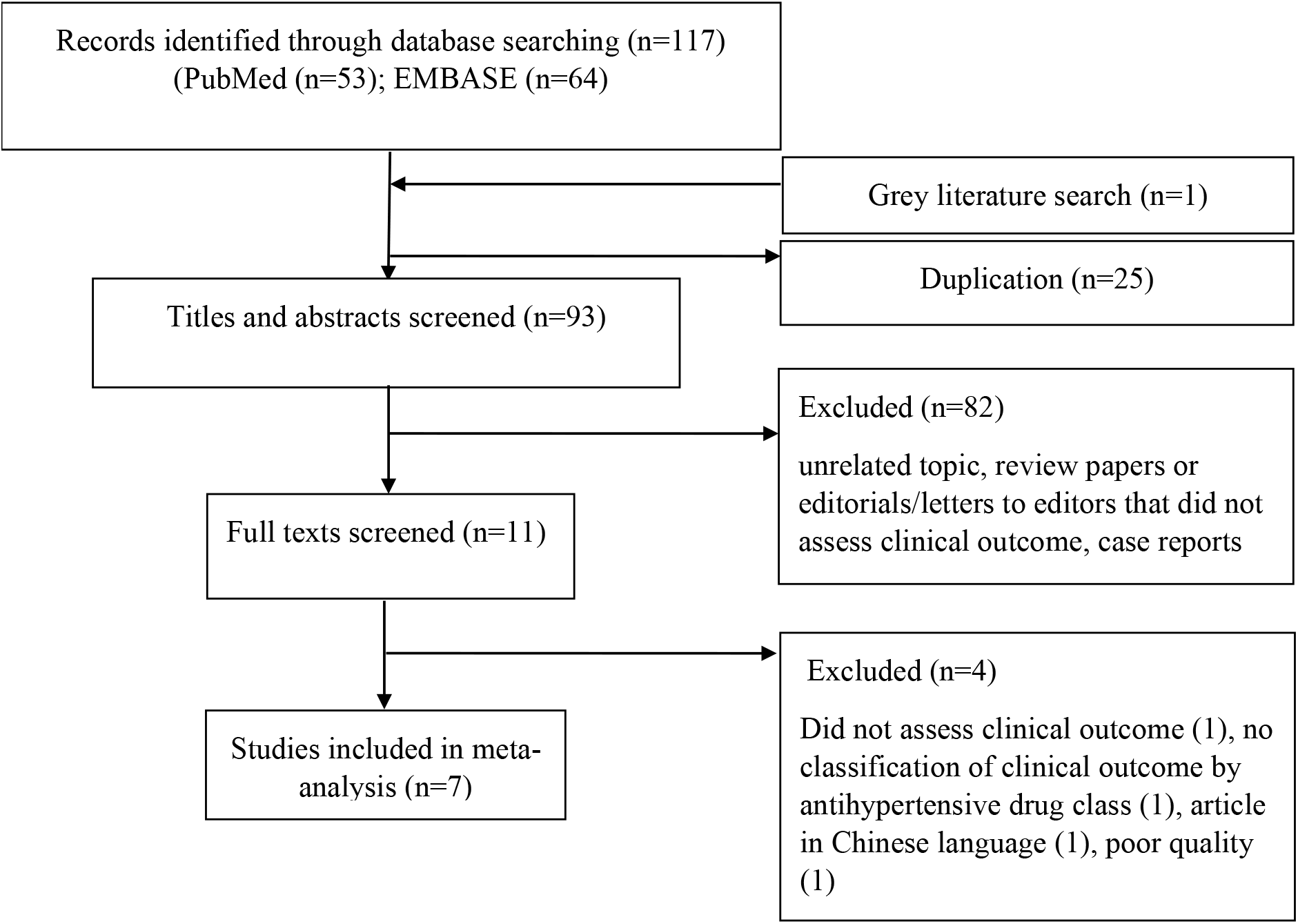
Flow chart showing the selection of articles for the meta-analysis.

**Table 1:**
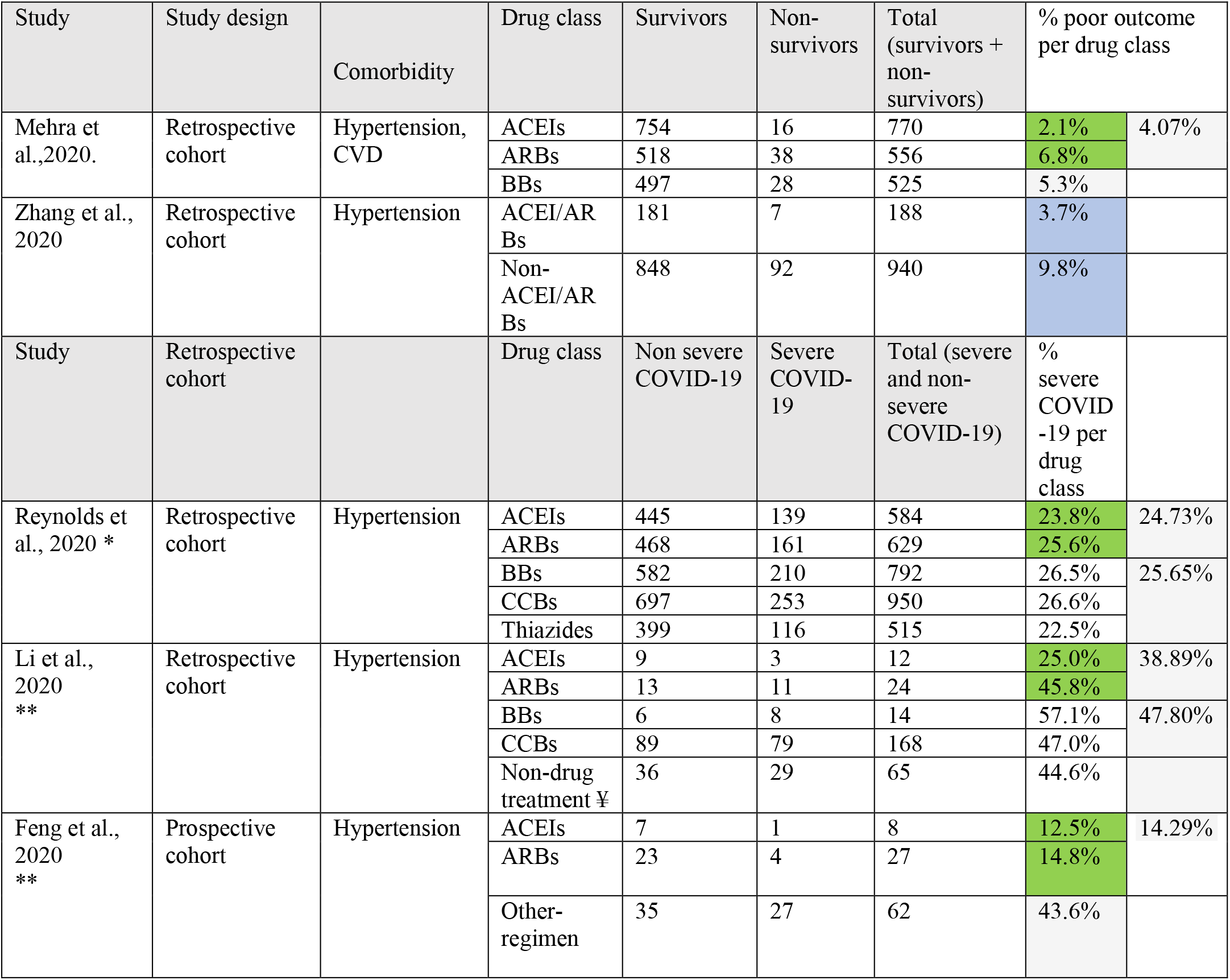
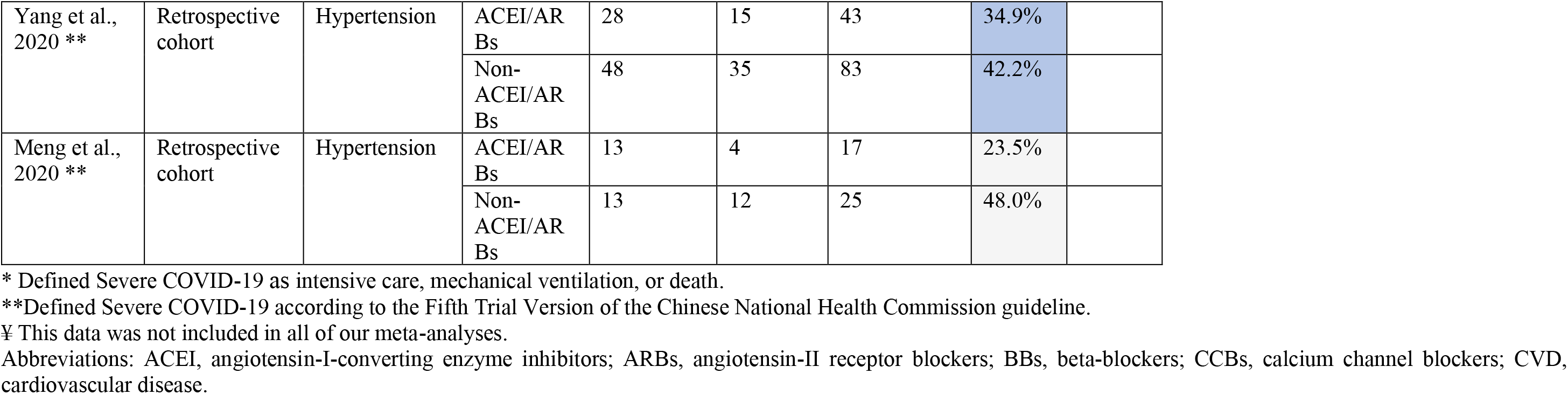
General characteristics of enrolled patients.

A total of 6,997 COVID-19 patients were included, and all of them had hypertension. The majority (58% or 4074/6,997) were taking non-RAAS inhibitors, whereas 41% (2,858/6,997) were receiving RAAS inhibitors. The remaining 1% were on non-drug treatment (included for descriptive purposes only and not included in the meta-analyses) for their hypertension. Most of the studies (5 of the 7 studies)^5,8–11^ categorized patients based on severity of COVID-19, of which 28% patients (1,107/4,018) developed severe COVID-19. The remaining two studies^6,7^ classified patients by survival status, and 6% (181/2,979) died. Only four studies^5,7–9^ documented the number of patients taking the specific drug class within the RAAS inhibitor and non-RAAS inhibitor groups. In these studies, the total number of patients taking ACEIs (1374), ARBs (1236), CCBs (1118) and BBs (1331) were approximately comparable (Table 1). The percentage of poor COVID-19 outcomes by class was lower with ACEIs compared with ARBs in all of these studies (Table 1).

### Comparison of the risk of poor COVID-19 clinical outcomes with different antihypertensives

We found that the overall risk of poor patient outcomes was lower in patients taking RAAS inhibitors (OR=0.84, 95% CI: [0.73, 0.96]; P=0.017) compared with those taking non-RAAS inhibitors. Poor clinical outcomes were less likely in patients receiving ACEIs (OR=0.73, 95% CI: [0.58-0.92]; P=0.01) than those receiving ARBs (Table 2). Compared to patients taking all other classes of antihypertensives, patients receiving ACEIs were less likely to experience poor outcomes (OR=0.77, 95% CI: [0.63-0.93]; P=0.002). However, we did not find a decreased risk of poor clinical outcomes in patients taking ARBs compared with those receiving all other antihypertensives (OR=1.13, 95% CI: [0.95-1.35]; P=0.02) (Table 2). The occurrence of poor patient outcomes was significantly lower in patients taking ACEs than those on BBs (OR=0.75, 95% CI: [0.60-0.95]; P= 0.02). There was no statistically significant difference in clinical outcomes between patients taking CCBs and ACEIs (Tables 1 and 2

**Table 2:** Risk of poor COVID-19 clinical outcomes with different classes of antihypertensives.

| Comparision | Odds ratio (meta-analysis) | 95% CI | P-value | Forest plot |
| --- | --- | --- | --- | --- |
| ACEI to ARBs | 0.73 | 0.58-0.92 | **0.01** | Supp. Figure S1 |
| ACEIs to BBs | 0.75 | 0.60-0.95 | **0.02** | Supp. Figure S2 |
| ACEIs to CCBs | 0.84 | 0.66-1.06 | 0.84 | Supp. Figure S3 |
| ACEIs to all other antihypertensives | 0.77 | 0.63-0.93 | **0.002** | Supp. Figure S4 |
| ACEIs to Non-drug treatment | 0.4* | 0.1-0.7 |  |  |
| ARBs to BBs | 1.0 | 0.81-1.23 | 0.44 | Supp. Figure S5 |
| ARBs to CCBs | 0.95 | 0.94-0.96 | - | Supp. Figure S6 |
| ARBs to all other antihypertensives | 1.13 | 0.95-1.35 | **0.02** | Supp. Figure S7 |
| CCBs to ACEI, ARBs, BBs | 1.07 | 0.90-1.27 | 0.86 | Supp. Figure S8 |
| ACEI, ARBs, BBs to CCBs and thiazides | 1.01 | 0.87-1.17 | 0.69 | Supp. Figure S9 |

\*Odds ratio only from one study (Li et al., 2020)
Abbreviations: ACEI, angiotensin-I-converting enzyme inhibitors; ARBs, angiotensin II receptor blockers; BBs, Beta blockers; CCBs, calcium channel blockers, Supp., supplementary.

## Discussion

Evidence on the safety of antihypertensive medications is of paramount importance as about one-third of the world’s population is estimated to have hypertension^18^ and this comorbidity is associated with increased mortality in patients with COVID-19^19^. Since RAAS inhibitors were reported to affect the clinical outcome of COVID-19 either for good or worse^6,12,20^, we pooled the recent studies to provide stronger evidence on the effects of these drugs. In addition, we also performed sub-meta-analyses to identify drug classes associated with better outcomes. We found that COVID-19 patients taking RAAS inhibitors had an overall decreased risk of poor outcomes compared to those receiving non-RAAS inhibitors. Interestingly, we also found that only the ACEIs (and not ARBs) were associated with decreased risk of poor patient outcomes (Table 2).

The decreased risk of COVID-19 severity or mortality with the use of RAAS inhibitors could be related to the blockage of a rapidly progressing systemic inflammation that is frequently seen in severe COVID-19^21^. For example, COVID-19 patients taking ACE/ARBs had lower levels of inflammatory markers, such as interleukin 6 (IL-6)^10^, C-reactive protein (CRP) and procalcitonin^11^, than those not taking these drugs. In addition, both classes of drugs could also help prevent hypokalaemia, a complication that was reported to occur in COVID-19 patients^22^. Hence, RAAS inhibitors may decrease poor clinical outcomes by limiting the deleterious effects of angiotensin-II in multisystem inflammation, as well as by preventing the occurrence of hypokalaemia^21,22^. However, it should be noted that another study found that the use of ACEIs/ARBs (versus no use) was associated with higher mortality in COVID-19 patients with cardiovascular disease^12^. This study was excluded from our meta-analysis as it involved a large number of patients with myocardial damage. Regardless, we also performed a separate analysis which included this study, and still poor COVID-19 clinical outcomes were lower with the use of RAAS inhibitors (OR=0.85, 95% CI: [0.74, 0.97]; P=0.016) (Supplementary Figure S10).

Further sub-analysis suggested that only ACEIs (and not ARBs) decreased the risk of poor clinical outcomes in COVID-19. This could be due to the differing mechanism of action of these drugs. While ACEIs block the synthesis of angiotensin-II, ARBs only block its action in certain receptors and do not affect its synthesis. Therefore, a high level of angiotensin-II in blood and tissues still occurs with the use of ARBs (versus low levels with ACEIs)^23^. It is known that there are different kinds of receptors for angiotensin-II, and the angiotensin-II receptor 1 (AT1R) is mainly blocked by ARBs. Hence, ARBs may not block the inflammatory effects of angiotensin-II mediated through receptors other than AT1R. Animal studies have shown that in order to fully stop the inflammatory effects of angiotensin-II, a combined blockade of both AT1 and AT2 receptors, as well as inhibition of its effects through the nuclear factor-κB (NF-κB) pathway, are required^24^. Therefore, our results do not support the claims that ARBs could have a therapeutic role in COVID-19^25^. However, ACEIs warrant further study as a potential repurposed drug therapy for COVID-19.

This meta-analysis has several limitations. First, we included a small number of studies that may affect the power of our conclusions. Second, even though all of the included papers were of good quality, propensity matching to address common confounders was performed in only two studies. In addition, there could also be a possibility of publication bias as all odds ratios were less than one (Figure 1), giving an asymmetry in the funnel plot. The other intrinsic problem of comparing one drug therapy to the other is that the indication of a certain antihypertensive usually depends on the presence of another coexisting comorbidity. For example, a comparison of RAAS inhibitors with non-RAAS inhibitors such as BBs might give a false sense of good outcome with RAAS blockers. This is because BBs might be commonly indicated as a rate controller for arrythmias that directly affect the prognosis. However, ACEIs and ARBs have fairly similar indications and our comparison of the risk between these two drugs should be more reliable.

**Figure 1:**
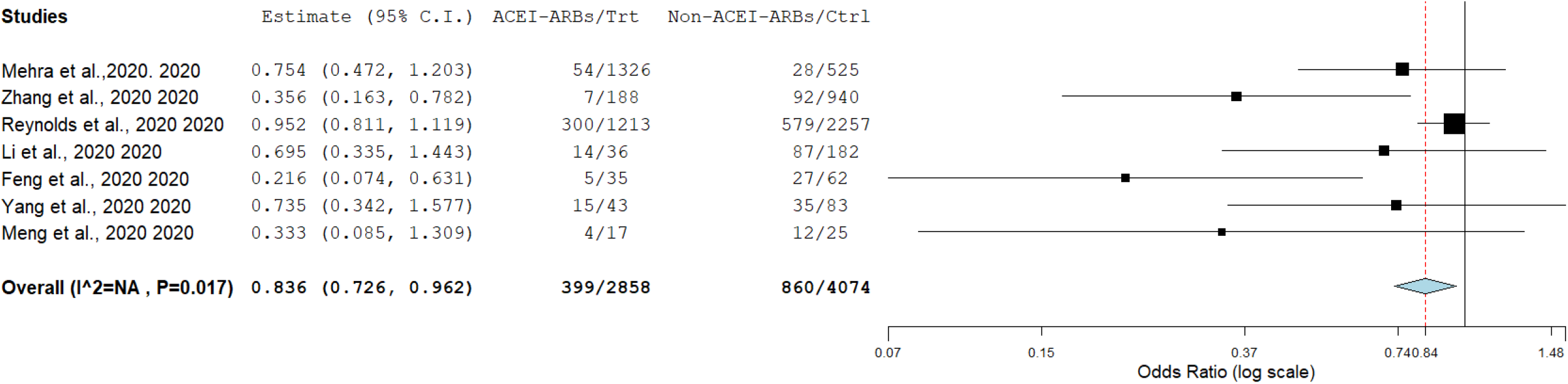
The risk of poor COVID-19 clinical outcome with ACEI/ARBS compared to Non-ACEI/ARBs.

In conclusion, the risk of severe COVID-19 or death was less likely in patients receiving RAAS inhibitors compared to those taking non-RAAS inhibitor antihypertensive agents. ACEIs could potentially decrease the severity and mortality of COVID-19.

## Data Availability

All data generated or analysed during this study are included in this published article.

## Author Contributions

Conceptualization, Y.B. and W.B.; methodology, Y.B. and W.B.; validation, Y.B., G.P., W.B, E.A. and A.B.; formal analysis, Y.B.; investigation, Y.B., W.B.; data curation, Y.B. and W.B.; writing—original draft preparation, Y.B.; writing—review and editing, Y.B., G.P., E.A., A.B.; visualization, Y.B., G.P., E.A., and A.B.; supervision, W.B.; project administration, W.B. and Y.B. All authors have read and agreed to the published version of the manuscript.

## Acknowledgments

Special thanks to Yohannes Gishen for his continuous support in this work.

## Annexes

**Supplementary Table 1:** Quality score of articles (Newcastle–Ottawa Scale)

|  | Selection (representativeness of exposed cohort, selection of the non-exposed cohort, ascertainment of exposure, at the start of the study the outcome of interest was not present) |  |  |  | Comparability (study design and analysis, and whether any confounding variables were adjusted) | Outcome (follow-up period, cohort retention, ascertained by independent blind assessment, record linkage, or self-report) |  |  |  |
| --- | --- | --- | --- | --- | --- | --- | --- | --- | --- |
| Study | Representativeness of Exposed Cohort (max: **) | Selection of the Non-Exposed Cohort from Same Source as Exposed Cohort: (*) | Ascertainment of Exposure (**) | Outcome of Interest Was Not Present at Start of Study (yes=*) | Comparability of Cohorts (**) | Assessment Outcome (**) | Follow-Up Long Enough for Outcome to Occur (*) | Adequacy of Follow-Up (**) | Quality Score |
| Reynolds et al., 2020 | ★★ | Yes ★ | Yes ★ | No | Yes ★ ★ | ★ | ★ | ★ | Good |
| Yang et al., 2020 | ★ | Yes ★ | Yes ★ | No | Yes ★ | ★ | ★ | ★ | Good |
| Li et al., 2020 | ★ | Yes ★ | Yes ★ | NO | Yes ★ | ★ | ★ | ★ | Good |
| Zhang et al., 2020 | ★★ | Yes ★ | ★ | Yes ★ | Yes ★ ★ | ★ | ★ | ★ | Good |
| Guo et al., 2020 | ★ | No | Yes ★ | No ★ | No | ★ | ★ | ★ | Poor |
| Meng et al., 2020 | ★ | Yes ★ | Yes ★ | No ★ | Yes | ★ | ★ | ★ | Good |
| Mehra et al.,2020. | ★★ | Yes ★ | Yes ★ | No ★ | Yes ★★ | ★ | ★ | ★ | Good |
| Feng et al., 2020 | ★ | Yes ★ | Yes ★ | No ★ | Yes ★ | ★ | ★ | ★ | Good |

Interpretation: Good quality: 3 or 4 stars (★) in selection domain AND 1 or 2 stars in comparability domain AND 2 or 3 stars in outcome domain; Fair quality: 2 stars in selection domain AND 1 or 2 stars in comparability domain AND 2 or 3 stars in outcome/exposure domain; Poor quality: 0 or 1 star in selection domain OR 0 stars in comparability domain OR 0 or 1 stars in outcome/exposure domain.

**Supplementary Figure S1:**
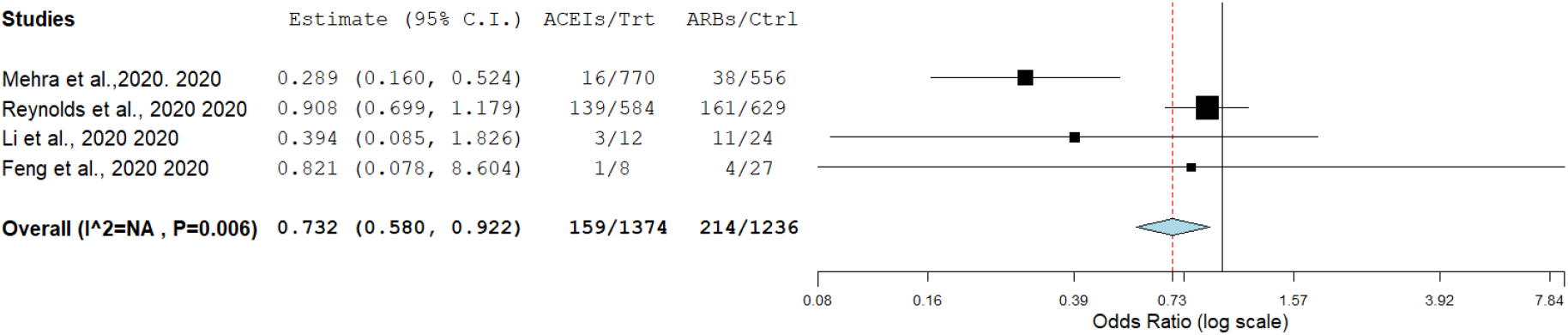
Risk of poor COVID-19 clinical outcome with ACEIs relative to ARBs.

**Supplementary Figure S2:**
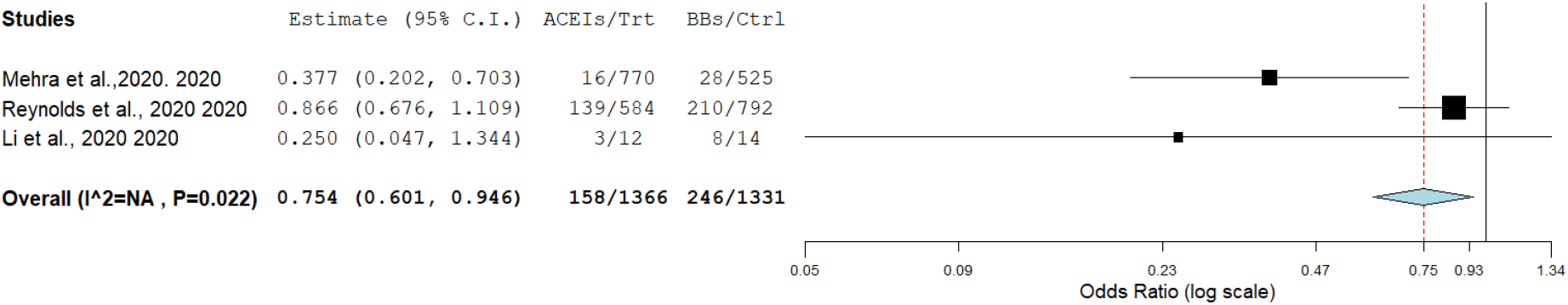
Risk of poor COVID-19 clinical outcome with ACEIs relative to BBs.

**Supplementary Figure S3:**
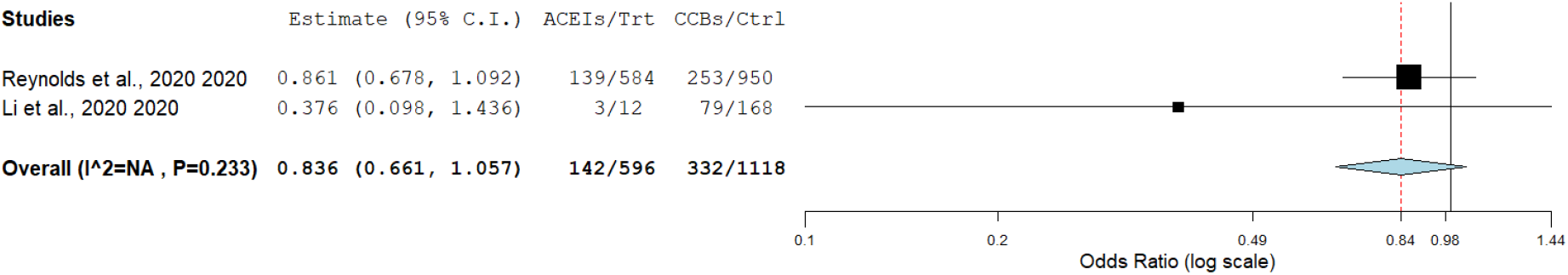
Risk of poor COVID-19 clinical outcome with ACEIs relative to CCBs.

**Supplementary Figure S4:**
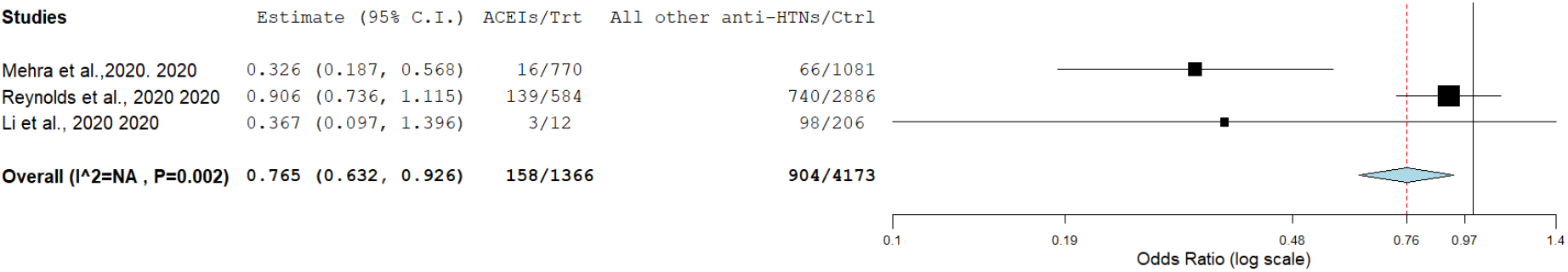
Risk of poor COVID-19 clinical outcome with ACEIs relative to all other antihypertensives.

**Supplementary Figure S5:**
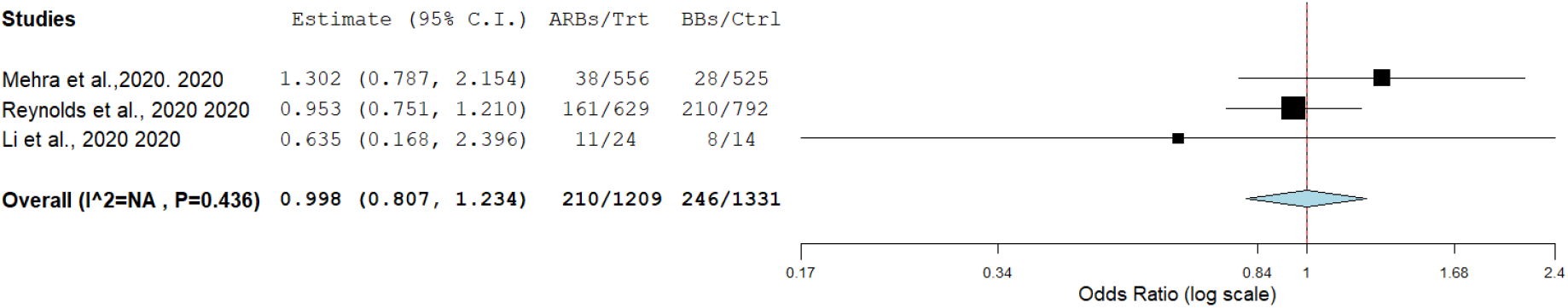
Risk of poor COVID-19 clinical outcome with ARBs relative to BBs.

**Supplementary Figure S6:**
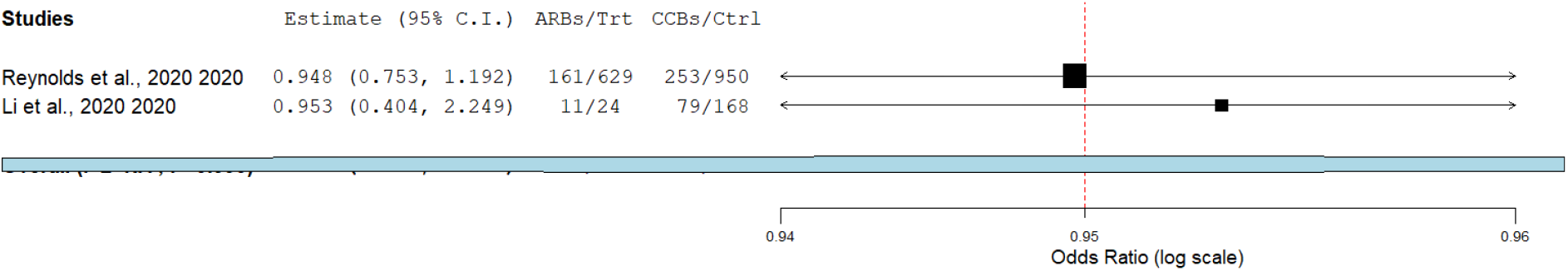
Risk of poor COVID-19 clinical outcome with ARBs relative to CCBs.

**Supplementary Figure S7:**
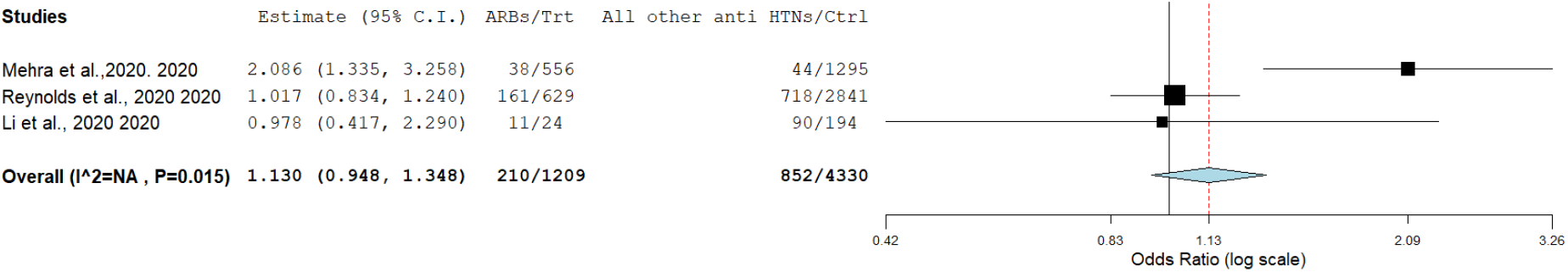
Risk of poor COVID-19 clinical outcome with ARBs relative to all other antihypertensives.

**Supplementary Figure S8:**
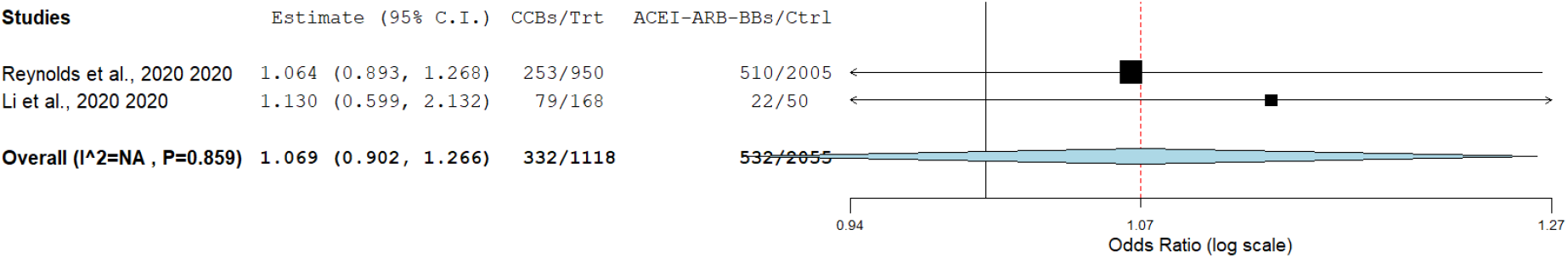
Risk of poor COVID-19 clinical outcome with CCBs relative to ACEI, ARBs, BBs.

**Supplementary Figure S9:**
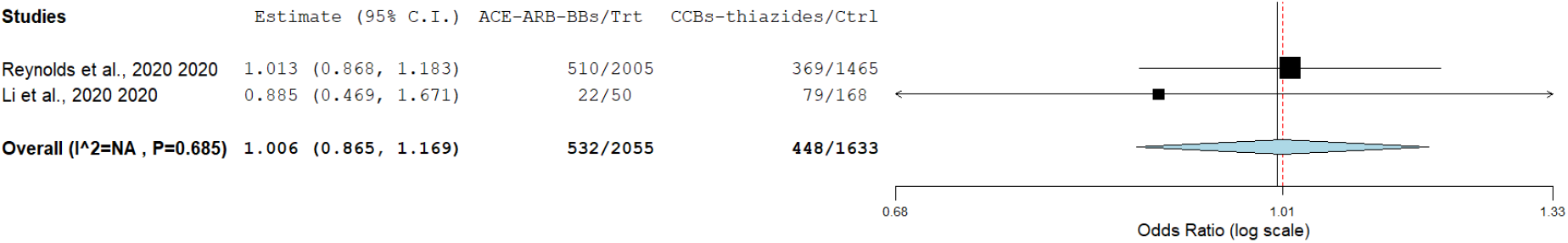
Risk of poor COVID-19 clinical outcome with ACEI, ARBs, BBs compared to CCBs and thiazides.

**Supplementary Figure S10:**
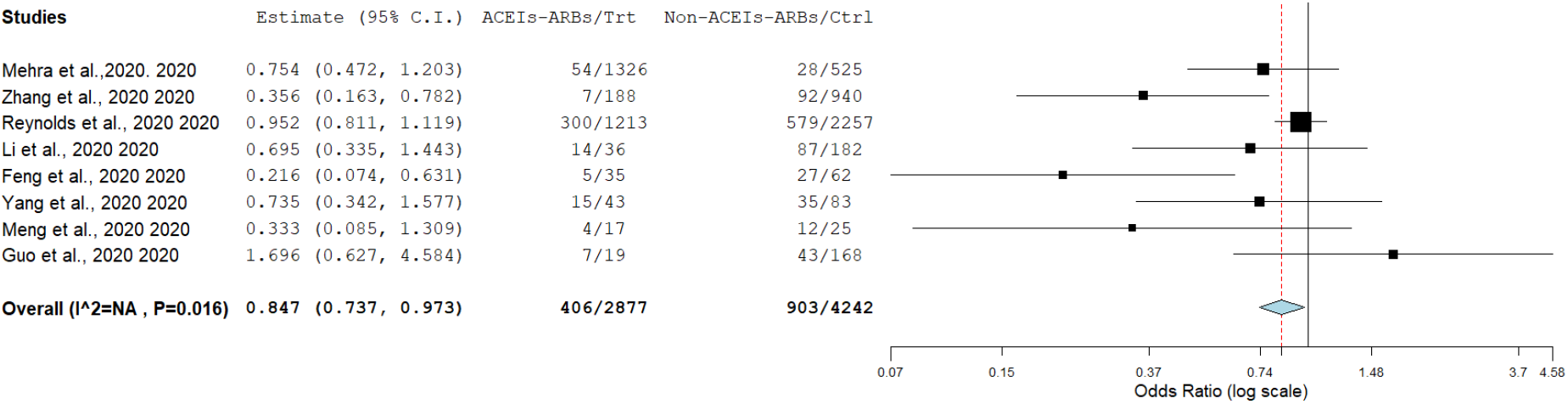
Risk of poor COVID-19 clinical outcome with ACEIs-ARBs compared to non-ACEIs-ARBs with Guo et al., 2020 added.

## References

1. Hanff, T. C., Harhay, M. O., Brown, T. S., Cohen, J. B. & Mohareb, A. M. Is There an Association Between COVID-19 Mortality and the Renin-Angiotensin System-a Call for Epidemiologic Investigations. Clin. Infect. Dis. Off. Publ. Infect. Dis. Soc. Am. (2020) doi:10.1093/cid/ciaa329.

2. Danser, A. H. J., Epstein, M. & Batlle, D. Renin-Angiotensin System Blockers and the COVID-19 Pandemic: At Present There Is No Evidence to Abandon Renin-Angiotensin System Blockers. Hypertens. Dallas Tex 1979 HYPERTENSIONAHA12015082 (2020) doi:10.1161/HYPERTENSIONAHA.120.15082.

3. Ferrario, C. M. et al. Effect of angiotensin-converting enzyme inhibition and angiotensin II receptor blockers on cardiac angiotensin-converting enzyme 2. Circulation 111, 2605-2610 (2005).

4. Hoffmann, M. et al. SARS-CoV-2 Cell Entry Depends on ACE_2_ and TMPRSS2 and Is Blocked by a Clinically Proven Protease Inhibitor. Cell S0092867420302294 (2020) doi:10.1016/j.cell.2020.02.052.

5. Feng, Y. et al. COVID-19 with Different Severity: A Multi-center Study of Clinical Features. Am. J. Respir. Crit. Care Med. rccm.202002–0445OC (2020) doi:10.1164/rccm.202002-0445OC.

6. Zhang, P. et al. Association of Inpatient Use of Angiotensin Converting Enzyme Inhibitors and Angiotensin II Receptor Blockers with Mortality Among Patients With Hypertension Hospitalized With COVID-19. Circ. Res. CIRCRESAHA.120.317134 (2020) doi:10.1161/CIRCRESAHA.120.317134.

7. Mehra, M. R., Desai, S. S., Kuy, S., Henry, T. D. & Patel, A. N. Cardiovascular Disease, Drug Therapy, and Mortality in Covid-19. N. Engl. J. Med. NEJMoa2007621 (2020) doi:10.1056/NEJMoa2007621.

8. Reynolds, H. R. et al. Renin–Angiotensin–Aldosterone System Inhibitors and Risk of Covid-19. N. Engl. J. Med. NEJMoa2008975 (2020) doi:10.1056/NEJMoa2008975.

9. Li, J., Wang, X., Chen, J., Zhang, H. & Deng, A. Association of Renin-Angiotensin System Inhibitors With Severity or Risk of Death in Patients With Hypertension Hospitalized for Coronavirus Disease 2019 (COVID-19) Infection in Wuhan, China. JAMA Cardiol. (2020) doi:10.1001/jamacardio.2020.1624.

10. Meng, J. et al. Renin-angiotensin system inhibitors improve the clinical outcomes of COVID-19 patients with hypertension. Emerg. Microbes Infect. 9, 757–760 (2020).

11. Yang, G. et al. Effects Of ARBs And ACEIs On Virus Infection, Inflammatory Status And Clinical Outcomes In COVID-19 Patients With Hypertension: A Single Center Retrospective Study. Hypertension HYPERTENSIONAHA.120.15143 (2020) doi:10.1161/HYPERTENSIONAHA.120.15143.

12. Guo, T. et al. Cardiovascular Implications of Fatal Outcomes of Patients With Coronavirus Disease 2019 (COVID-19). JAMA Cardiol. (2020) doi:10.1001/jamacardio.2020.1017.

13. PRISMA. http://prisma-statement.org/prismastatement/Checklist.aspx.

14. [Interpretation of “Guidelines for the Diagnosis and Treatment of Novel Coronavirus (2019-nCoV) Infection by the National Health Commission (Trial... - PubMed - NCBI. https://www.ncbi.nlm.nih.gov/pubmed/32033513.

15. Shah, A., Gandhi, D., Srivastava, S., Shah, K. J. & Mansukhani, R. Heart Failure: A Class Review of Pharmacotherapy. Pharm. Ther. 42, 464–472 (2017).

16. Wells, G. et al. Ottawa Hospital Research Institute. http://www.ohri.ca/programs/clinical_epidemiology/oxford.asp.

17. Wallace, B. C. et al. *Open MEE*: Intuitive, open-source software for meta-analysis in ecology and evolutionary biology. Methods Ecol. Evol. 8, 941–947 (2017).

18. Mills, K. T., Stefanescu, A. & He, J. The global epidemiology of hypertension. Nat. Rev. Nephrol. 16, 223–237 (2020).

19. Zuin, M. et al. Arterial hypertension and risk of death in patients with COVID-19 infection: Systematic review and meta-analysis. J. Infect. S0163445320301894 (2020) doi:10.1016/j.jinf.2020.03.059.

20. COVID-19 and the cardiovascular system: implications for risk assessment, diagnosis, and treatment options | Cardiovascular Research | Oxford Academic. https://academic.oup.com/cardiovascres/advance-article/doi/10.1093/cvr/cvaa106/5826160.

21. [Inhibitors of RAS Might Be a Good Choice for the Therapy of COVID-19 Pneumonia] - PubMed. https://pubmed.ncbi.nlm.nih.gov/32061198/.

22. Hypokalemia and Clinical Implications in Patients with Coronavirus Disease 2019 (COVID-19) | medRxiv. https://www.medrxiv.org/content/10.1101/2020.02.27.20028530v1.

23. Bomback, A. S. & Toto, R. Dual Blockade of the Renin–Angiotensin–Aldosterone System: Beyond the ACE Inhibitor and Angiotensin-II Receptor Blocker Combination. Am. J. Hypertens. 22, 1032-1040 (2009).

24. Ruiz-Ortega, M. et al. Angiotensin II: a key factor in the inflammatory and fibrotic response in kidney diseases. Nephrol. Dial. Transplant. 21, 16–20 (2006).

25. Network-based drug repurposing for novel coronavirus 2019-nCoV/SARS-CoV-2 | Cell Discovery. https://www.nature.com/articles/s41421-020-0153-3.

